# Harms from Heat-Health Risks: Morbidity Evidence from India and Global Learnings for Policy Action

**DOI:** 10.1101/2025.07.07.25330912

**Authors:** Purnamita Dasgupta, Rajib Dasgupta, Kristie L Ebi, Girika Sharma, Madhura Chowdhuri, Shivangi Shankar, Shreya Pujari

## Abstract

Increasing temperatures in India, along with a rise in the frequency, intensity and duration of heatwaves, pose health risks. Similar challenges from heat events are being faced across geographies, in both high and low-and-middle-income countries. This paper examines the evidence on heat-health risks for the Indian population using a national level dataset on over 500,000 individuals and 63 illnesses. It also synthesizes global evidence and scholarship on heat-health risks through a narrative review of the literature on morbidity outcomes. The results from the data analysis and the synthesized findings establish that significant morbidity is associated with heat stress experienced during the summer months, and that specific illnesses are aggravated by heat, especially for vulnerable sub-groups such as older adults, females and outdoor workers. The paper presents evidence on stressors and factors that influence health outcomes including pre-existing illnesses, socio-economic vulnerability and planned heat adaptation responses. It identifies the determinants of risk, specific knowledge gaps for further research and multiple options for risk management which can be considered particularly for low income contexts and LMICs, including India. The study provides an evidence base along with specific recommendations to inform policy on adoption of short- and long-term strategies for reducing HRIs and strengthening health system resilience. There is an urgent need for health sector actors to actively engage in expanding evidence on heat related health risks and in building resilience.

## 1. INTRODUCTION

Key international conventions that focus on achieving resilient and sustainable communities recognize that good health is essential for sustainable development [1, 2]. The impacts of climate change on human health are severe and already being felt globally [3, 4]. The global urgency for the convergence of climate science, technology, and public health to support health and well-being is well-established [5]. The World Health Organization (WHO) has been strengthening its engagement with agencies such as the Intergovernmental Panel on Climate Change (IPCC), the World Meteorological Organization (WMO) and United Nations Development Programme.

A major challenge for many countries is the mortality and morbidity from high heat [6, 7]. These have economic consequences, such as reduction in the number of hours worked due to heatwaves and job losses, declining productivity, damage to health infrastructure and increased health costs. Investments in cost-effective interventions can increase resilience for these largely preventable heat-health risks [8]. This paper adds evidence to support the urgent call for a health-centered approach [9] towards risk reduction and effective management of heat stress, contributing to the advancement of the sustainable development goal on health. 2024 was the warmest year in 175 years of observational record. The rising number of heat wave days and heat exposure poses record-breaking threats to health and well-being, especially for the vulnerable, such as older adults [10]. Evidence establishes that temperatures are increasing in India, along with a rise in the frequency, intensity and duration of heatwaves, increasing risks to human health [11, 12, 13, 14]. Models project that heat events would continue to increase in the future [15]. Heat stress is associated with heat rash, cramps, exhaustion, heat stroke, hospitalisation, and deaths from respiratory, cardiovascular and renal diseases. Vulnerability to heat is a major concern for India given the presence of a population of more than 1.4 billion, with a little less than half of the urban population residing in low income settlements, and a similar proportion of workers employed outdoors.

Several studies provide evidence for all cause- and cause-specific mortality attributable to heat in various regions and cities ( Supplementary Material, Table 1, SM1 provides details). There is a dearth of empirical studies that examine morbidity and the pathways through which health risks can be lowered, especially in LMIC contexts. This paper presents statistical evidence from India on seasonal variations in occurrence of non-communicable diseases (NCDs). Subsequently, a narrative review of the literature on heat-health stressors is used to identify options for risk reduction and knowledge gaps in the understanding of heat-health risks in LMICs.

**Table 1.** Classification by availability and strength of evidence.

| STAGE 1: Availability of evidence |  |  |  |
| --- | --- | --- | --- |
|  | HIC | LMIC (Excluding India) | India |
| Established | At least two studies | At least one study | At least two studies |
| Not Established | Less than two studies | No studies | Less than two studies |
| **STAGE 2: Availability and strength of evidence** |  |  |  |
|  | **HIC** | **LMIC (Excluding India)** | **India** |
| Well established | More than two studies + Similar Findings | More than two studies + Similar Findings | More than four studies + Similar Findings |
| Partially Established | Two studies + Similar Findings | One to two studies + Similar Findings | Two studies + Similar Findings |
| Not Established | Less than two studies or Dissimilar findings* | No studies or Dissimilar findings | Less than two studies or Dissimilar findings |

Notes: 1. In case of establishing global evidence, both HIC and LMIC conditions have to be simultaneously fulfilled.
2. In instances where there were dissimilarities in the findings in terms of the directionality of the impacts of heat on human health irrespective of whether this occurred in an LMIC or HIC context, the evidence for the corresponding risk factor was considered as not established. This holds even if a single study arrived at a contrary or ambiguous finding on the corresponding risk factor.
3. Textual description available in SM3.

### Objectives

Observational evidence on the morbidity impacts of extreme heat and heatwaves in LMICs is rare. Measures to reduce heat risks tend to draw upon observational evidence from the Global North and physiological understanding. Variations in bio-meteorological, climatic, and social determinants of health call for contextualised evidence for accurate identification of risk as well as the management and prevention of health effects in countries like India [15, 16]. As heat action plans are being devised and implemented in parts of India, it is important to examine closely the factors that affect heat-health risks [17]. There are significant advances that have been made in the global scholarship on how heat affects health. These are relevant for risk management in LMICs, when complemented with contextualized learnings.

The paper has two objectives. Firstly, to provide empirical evidence on the substantial morbidity risks associated with heat stress, establishing the urgent need to expand the evidence base. Secondly, to synthesize global learnings on heat-health risks, identifying the determinants, knowledge gaps and multiple options for risk management in low income contexts, using evidence and examples from India.

## 2. METHODOLOGY

Objective 1 is addressed through analyzing evidence on morbidity, using the publicly available National Sample Survey (NSS) dataset. Objective 2 is pursued through an analytical synthesis of the literature on the determinants of heat-health risks.

### Objective 1: Evidence from morbidity reporting

The 75th round of survey by the National Sample Survey (NSS 2019) conducted from July 2017 to June 2018 was used. The dataset provides information at the household level on morbidity, socio economics, demographic, and health expenditure domains from household members. The nation-wide survey covered over a hundred thousand (113,823) households and five hundred thousand (555,352) individuals and captured details on socio-economic and demographic characteristics, including self-reported illnesses, health care seeking behaviour, and health expenditures. Information was available on 63 illnesses, classified under 15 broad categories including Cardiovascular, Respiratory, and Infectious diseases. Respondents provided information on the occurrence of illnesses during the 15 days immediately preceding the date of the survey. Because the survey date is available, each individual’s reporting of one or more illness episodes in the preceding 15 days could be mapped onto the month (and weather) during which the illness had occurred, alongside the individual’s demographic details. 43, 219 individuals reported data on illnesses.

District-level monthly temperature data (ICRISAT 2025, details in SM2) on highest maximum and lowest minimum temperatures for all districts across India was also analysed for 2017-2018 Figure 1 provides the distribution of the districts in terms of the highest monthly maximum temperatures recorded, across the twelve months in 2017-18, the year corresponding to the illness survey. The data on temperatures was binned for ease of representation.

**Figure1.**
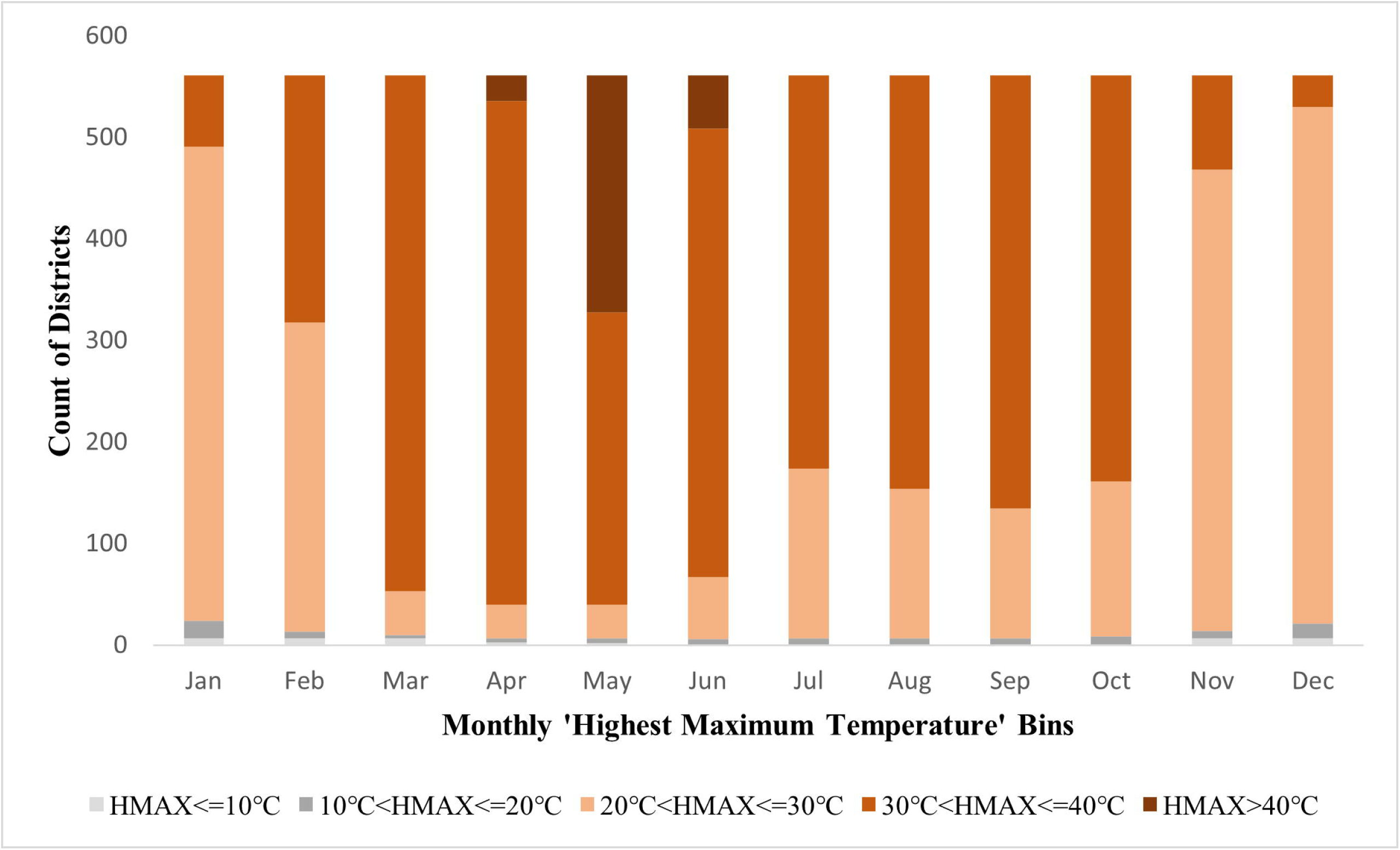
Distribution of 561 Indian Districts by Monthly ‘Highest Maximum Temperature (HMAX)’ Bins.

India is a large country with significant climatic variations. However, between March to June, almost all states experience the summer season. March, April, May, and June experience predominantly higher temperatures across almost all the districts, with temperatures increasing beyond 30 degrees C in over 87% districts in March, thereafter reducing gradually from July onwards. Temperatures soar over 40°C by May in over 40% of the districts. A summer season is clearly identifiable and confirmed during the months of March to June. A similar pattern is observed in the minimum temperatures ( Figure 1, SM 2), with the lowest minimum temperature increasing from March onwards, peaking in June and declining from July onwards for almost all districts. A t-test comparing the means of the summer and non-summer months across districts yields a significant difference at 1 percent level (Table 1, SM 2).

Therefore, illness data from March to June (4 months) were considered for the summer months, and compared with data from the rest of the year, the latter being categorized as non-summer months. Descriptive statistics were used for mapping the differentials in reporting of illnesses during the summer versus the non-summer months. The associations between seasonality and the self-reported occurrence of illnesses were examined using the chi-square test. The test was set up to check whether there was a difference in self-reporting for illnesses in summer as compared to non-summer months. The null hypothesis was there was no difference, (H_O: no difference) and the alternative hypothesis that a difference existed (H_1: difference exists). Subsequently, odds ratios were calculated to determine the specific nature of the relationship, whether illness reporting was higher in summer months or non-summer months. Age, gender, education level, and dummy variables to capture state-specific factors were included as additional explanatory factors in the estimation.

The same approach was repeated for three sub-populations considered to be particularly vulnerable: older adults defined as aged 60 and above; female population; and those routinely engaged in outdoor activities for their occupation. This analysis was done for two specific illnesses—hypertension and diabetes--which had a high prevalence within the sample.

### Objective 2: Narrative Review

The narrative review was shaped by multiple perspectives of heat-health, from diverse domains of human physiology, clinical medicine with its sub-disciplines, public health, environmental health and environmental-health economics. Knowledge and models from diverse sources were pooled to enable the synthesis. A non-comprehensive search captured and identified dominant themes. There were no strict predetermined inclusion or exclusion criteria; the focus was on identifying relevant literature, assessed by the domain expertise of research team members. The search included electronic the database sources PubMed, MedInd, and Google Scholar. Given the evolving nature of the topic, additional searches were undertaken for technical reports, conference proceedings and similar materials [18, 19]. The researcher-team sought to ensure ‘meaning saturation’ – that all issues were fully understood till no new information about the meaning of themes and their relationships emerged [20].

The available evidence on the factors determining heat-related health risks was categorized as well-established, partially established, or not established based on a 2 stage review process, which assessed evidence in terms of the extent of coverage and strength of evidence. The extent of coverage considers the availability of evidence from low-and middle-income countries (LMICs) as well as high-income countries (HICs). The strength of evidence considers the extent to which the findings from available studies were similar, in terms of the directionality of the impacts of heat on health. At the first stage, availability of evidence from at least one LMIC country, apart from India, was considered as an essential criterion. If this criterion was unfulfilled, the global evidence on the corresponding risk factor was classified as not established. At the second stage, the strength of evidence on the risk factor was additionally considered. Table 1 presents the classification criteria.

## 3. FINDINGS ON MORBIDITY FROM THE SURVEY DATA ANALYSIS

The data (details in Table 1, SM4) is almost equally divided between male and female respondents, with a little less than half engaged in outdoor occupations. Occupations classified under the indoor category include those who reported that they were self-employed in household enterprises as either workers or employers, were unpaid helpers in household enterprises, worked as a regular salaried or wage employee, and those who did not work but were seeking or available for work. It also includes those attending domestic duties, or unable to work for some reason, and rentiers, pensioners, and remittance recipients. Outdoor occupations include those who work as casual wage labour in public works or other places, attend educational institutions, attend domestic duties along with major outdoor engagements such as collection of goods (vegetables, roots, firewood, cattle feed, etc.), and those employed outside the home in various sectors (for example, tailoring, weaving,).

Of the 42,762 older adults in the dataset, 30.1 percent were enumerated in summer months and 69.9 percent in non-summer months. 31.5% of the 272,115 females in the dataset were interviewed during the summer months, and 31.4% of the 242,581 individuals involved majorly in outdoor activities were surveyed in summer. The enumeration is well distributed across the months, supporting a cross-section analysis.

### Differentials in Reported Illnesses for all individuals surveyed

Out of the 63 self-reported illnesses in the dataset, 20 illnesses (32%) occurred more in summer than in the non-summer months. 17 percent of the illnesses tended to be reported equally across the two seasons. For the rest, the non-summer months had higher reporting, per thousand population (more details in Figure 1, SM4). The difference in reporting across summer and non-summer months varied substantially across illnesses, highest for hypertension (2.38), followed by diabetes (2.17), goitre and thyroid (0.58), heart disease (0.31), and bronchial asthma (0.16).

A chi-squared test for differences between the seasons was conducted for the 20 illnesses with higher reporting during the summer months (details in Table 2, SM 4). For hypertension, bleeding disorders, diabetes, goitre and thyroid, others including obesity and retardation, the test results are significant at the 5% level, and for heart disease at the 10% level. Odds ratios were calculated to examine the direction of reporting, using a logistic regression model that incorporated age, gender, education, and state dummies as additional independent variables (details in Table 3, SM 4). The odds ratios for six of these illnesses were significant at the 5% level, and for heart disease, at 10 % level.

**Table 2.** Results on Chi-squared test and odds ratio.

| **Older Population** |  |  |  |  |
| --- | --- | --- | --- | --- |
| **Illness** | **$\chi^2$ Value** | **p-Value** | **Odds ratio** | **p-Value** |
| Hypertension | 57.90 | 0.000** | 1.35 | 0.000** |
| Diabetes | 38.60 | 0.000** | 1.25 | 0.000** |
| **Female Population** |  |  |  |  |
| **Illness** | **$\chi^2$ Value** | **p-Value** | **Odds ratio** | **p-Value** |
| Hypertension | 38.07 | 0.000** | 1.28 | 0.000** |
| Diabetes | 20.54 | 0.000** | 1.19 | 0.000** |
| **Outdoor Workers** |  |  |  |  |
| **Illness** | **$\chi^2$ Value** | **p-Value** | **Odds ratio** | **p-Value** |
| Hypertension | 4.97 | 0.026** | 1.16 | 0.019** |
| Diabetes | 5.56 | 0.018** | 1.15 | 0.031** |

\*\* denotes significance at 5% alpha level ( $p < 0.05$ )

**Table 3.** Determinants of Morbidity Risks from Heat Stress.

| Determinants | Factors (stressors) | Global Evidence (HIC & LMIC) | India Evidence |
| --- | --- | --- | --- |
| Heat Hazard | Increasing Maximum Temperatures | Well established | Well established |
|  | Increasing Minimum Temperatures | Well established | Partially established |
|  | Rise in Humidity | Not established | Not Established |
|  | Rise in Heatwave Occurrence | Well established | Well established |
| Exposure | Outdoor work | Well established | Well established |
|  | Lack of ventilation | Well established | Partially Established |
|  | Indoor heat load | Well established | Well established |
| Biological/<br>Physiological<br>vulnerability | Age (Infants, Neonates, Older Adults) | Well established | Partially established |
|  | Pre-existing Illnesses (Chronic diseases e.g., cardiovascular, diabetes respiratory, kidney conditions, psychiatric conditions etc) | Well established | Not established |
|  | Short term occurrence of new symptoms (fatigue, exhaustion, fever) | Well established | Well established |
|  | Disorders of Sweating | Well established | Not Established |
|  | Medication | Well established | Not Established |
| Socio-economic<br>vulnerability | Income | Well established | Partially established |
|  | Housing Quality | Partially established | Partially established |
|  | Access to Cooling Resources (Energy, Equipment, Water) | Well established | Well established |
|  | Access to healthcare | Well established | Well established |
|  | Neighbourhood (Green and blue spaces) | Well established | Partially established |
| Sex & Gender<br>Vulnerability | Biological Differences | Not established | Not established |
|  | Sociocultural Factors | Partially established | Partially established |
|  | Access to Resources | Well established | Well established |
| Responses | Heat-Health Warning systems | Partially established | Partially established |
|  | Cooling Interventions | Well established | Well established |
|  | Public Awareness | Well established | Well established |
|  | Policy and Regulations | Well established | Well established |
|  | Synergistic Measures:<br>Urban heat island effect | Well Established | Partially established |
|  | Air Pollution | Not established | Not established |

### Differentials in Illnesses across vulnerable groups: Older Adults, Females, and Outdoor Workers

For probing illnesses amongst the vulnerable groups, the study considered illnesses reported by at least 1% of the surveyed population, namely 6369 for diabetes and 6563 for hypertension.

Amongst those reporting hypertension 54 % were older adults, 55% female and 19.2% outdoor workers; for diabetes, the corresponding numbers were 49% female, 17% outdoor workers, and 51% older adults (details in Table 4, SM 4). Figure 2 presents the break-up across summer and non-summer months.

**Figure 2.**
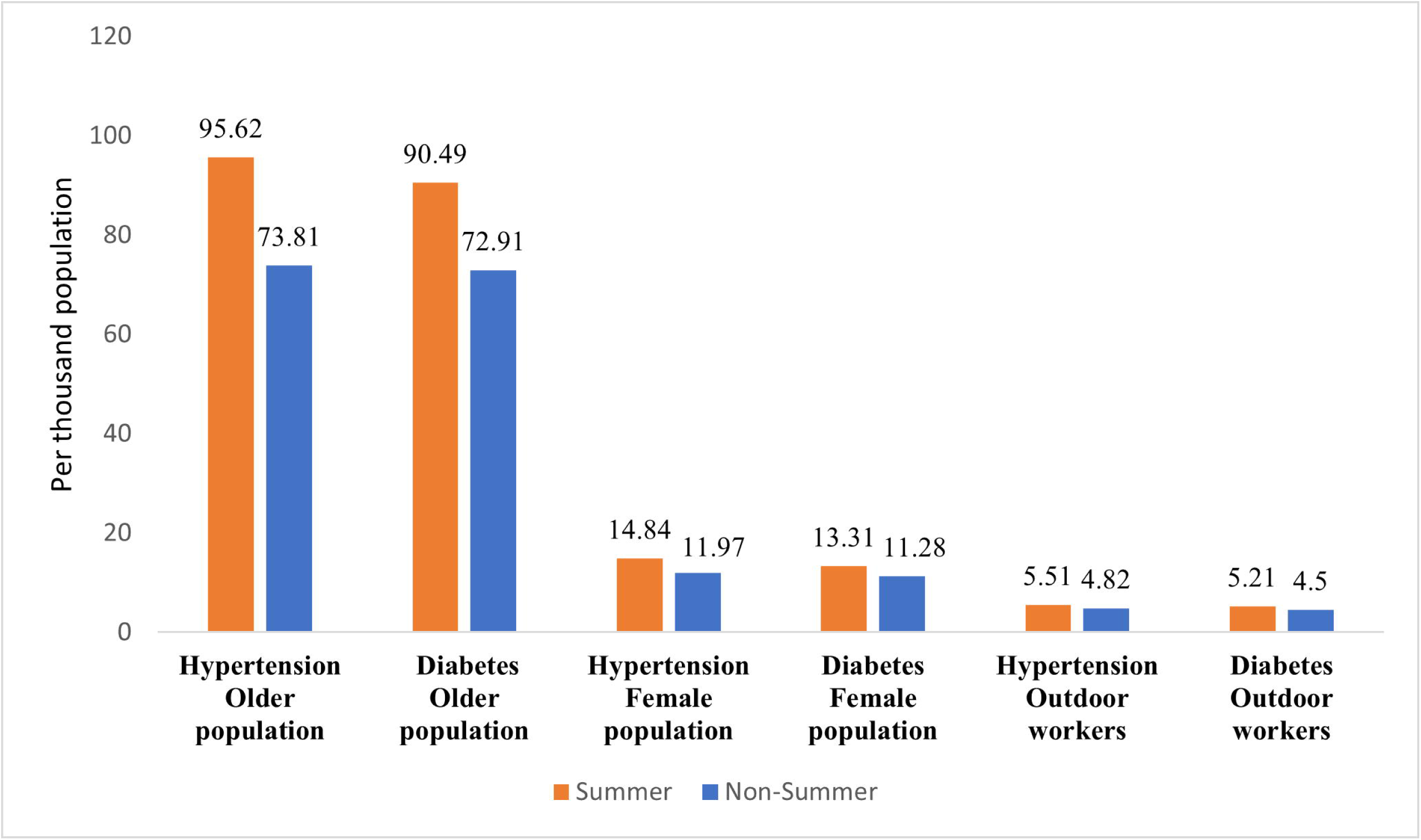
Diabetes and Hypertension reported during summer and non-summer months.

Chi-square tests indicated a significant difference in self-reported illnesses across seasons (summer and non-summer) at the 5% level of significance, for the three vulnerable categories. Additionally, odds ratios were calculated to explore the direction of reporting, after incorporating age, gender, education, and state dummy variables as explanatory factors. The odds ratios for both illnesses were significant at the 5% level and indicated that that self-reported cases were higher during the summer months for both hypertension and diabetes (see Table 2).

## 4. FINDINGS FROM THE HEAT-HEALTH REVIEW RELEVANT TO INDIA

### Heat Related Illnesses: Risk Framing

A risk framing for HRIs that targets the effective management of heat-health impacts must incorporate the diversity of factors that determine risks. Research on the physiological aspects of temperature regulation amongst workers in industrial and military settings began in the early 19^th^ Century [21]. The strain due to exposure to excess heat affects nearly all organ systems through multiple mechanisms [22], with higher risks amongst those suffering from pre-existing illnesses such as cardiovascular, kidney, and respiratory diseases, and mental health [23, 24].

Further, HRIs affect some population subgroups more than others due to factors such as availability of health care. Risks are compounded for LMICs with high non-communicable disease burdens, such as diabetes in India, [25, 22]. Older adults, infants, children, and pregnant women may also face higher risks of adverse impacts for biological and socio-cultural reasons in LMICs [26]. Socio-economic conditions, such as housing, neighbourhoods, workplace environments, and clothing practices, infrastructure and access to care have a major role in determining heat-health outcomes. Managing risks therefore calls for adequate and planned response options that can address the range of determinants of heat stress on human health [27].

The health risks of heat were accordingly framed in terms of its four determinants (stressors), in line with the framing of risks proposed by the Intergovernmental Panel on Climate Change [3]: (i) hazard (ii) exposure (iii) vulnerability and (iv) response, the latter being defined in terms of the inadequacy or absence of adaptation. Table 3 presents the major findings from the review, in alignment with this framing of heat-health risks. Column 1 presents the four determinants, column 2 expands on the specific factors within these broad determinants, and columns 3 and 4 present findings from the review on global evidence and the Indian evidence, respectively (more details in SM 5).

### Key learnings

Risk outcomes are influenced by a range of factors, and by the interactions between hazard, vulnerability, exposure, and the capacity of health systems to respond to high heat situations. It is possible to reduce morbidity and prevent deaths through effective risk management. Based on the evidence synthesis, a range of actions exist, including reducing air pollution, bringing down indoor heat loads, enhancing access to sustainable cooling, maximizing the co-benefits of nature-based solutions that promote green and blue spaces, enforcing work space regulations and enhancing the capacity of the health system. Much however, depends on the availability of reliable data and information.

The empirical evidence on heat-hazard risks from increases in maximum temperatures, minimum temperatures, and heat wave occurrences are well established, although health risks attributable to rising minimum temperatures in India needs strengthening. Evidence on health risks from humidity are not conclusively established, especially for LMICs, including India.

Exposure risks are mostly well-studied and understood. In LMIC contexts, evidence established the increased heat-health risks for outdoor workers in agriculture and construction, from workplace injuries in hotter months, and lack of ventilation. However, more studies are required on context-specific attribution particularly for densely populated, low-income, urban settlements with a high urban heat island impact. Indoor heat loads also increase risks, particularly from use of traditional stoves and biomass for cooking, and poor-quality housing.

Vulnerability is considered under three sub-categories, namely, biological, socio-economic, and gender-based vulnerability with the evidence grouped into 4-5 key domains. Established evidence on age-specific vulnerabilities focuses on infants, neonates, and older adults. Less is known about risks faced by other, school-going children. Heat-health risks could be high where children walk to school, have classrooms that are not air conditioned, and commute during peak heat hours. In India, children and elderly are a cause of concern, and more studies are required to enable attribution and risk management, with 138 million older adults and 265 million school children [28].

Heat-health risks are high for people with pre-existing illnesses, especially for diabetes, cardiovascular, respiratory, kidney, and psychiatric conditions. India specific studies are urgently required to help understand the causal pathways and strengthen responses. Mental illnesses are not sufficiently researched.

An increased risk of waterborne and vector borne diseases following exposure to heat waves and increased symptoms of fatigue, exhaustion, and fever were reported in multiple heat-health studies in LMICs. There is a lack of evidence on the health impacts of prolonged exposure to heat, interactions between high heat and humidity, on the adverse impacts of medications on thermoregulation, sweating capacities, and disorders, relevant to LMICs, including India.

Morbidity and mortality from HRIs are higher among those with lower incomes. Lack of income adversely impacts heat exposure and adaptation and increases the likelihood of employment in outdoor work. For India, studies on heat-related morbidity attribution for income, housing quality, and other socio-economic stressors is urgently required. Lack of access to healthcare and cooling options pose health risks across regions. It is well established that neighbourhood green and blue spaces are important for reducing heat-health risks, and much more can be done in India to assess and invest in planning such spaces.

Available empirical evidence from India or elsewhere, does not lead to a consensus on the extent to which sex determines susceptibility to mortality or morbidity outcomes due to heat. Bell et al. (2008) found that in Santiago, Chile and Brazil, mortality was higher among men, whereas women were more susceptible in Mexico City. Estimates vary for India [29]. Choudhary *et al*. (2023) reported higher female mortality in the cities of Delhi and Chennai, and higher male mortality in Varanasi [30]. Kumar and Singh (2021) reported higher mortality rates for men in several states in India [31]. Maternal and birth outcomes were impacted by heat in multiple ways, with studies reporting increased risks of near-fatal health incidents in pregnant mothers, higher incidence of preterm birth, low birth weights, and still-births [32, 33].

Socio-cultural practices impact heat - health risks differentially across communities and sub-groups, such as children and women. Such practices may relate to choice of clothing, place to sleep, and food and beverage consumption, access to housing, safe drinking water, toilets, cooling measures, communication technology, and early warning systems, with substantial evidence on differential access for women. With women more susceptible to urinary tract infections, outcomes can worsen with dehydration. A dearth of appropriate sanitation facilities in low-resource settings can lead to harmful practices such as drinking less water despite extreme heat to avoid urination.

Planned responses should reduce heat risks in an equitable and efficient manner. Evidence indicates it is possible to design and implement cooling interventions at individual, community, and healthcare delivery scales. Empirical evidence is strong on the potential for awareness creation and of policy and regulations as effective means of reducing heat-health risks. However, heat management plans, such as city level heat action plans (HAPs) and national health adaptation plans need to distinguish between risk factors that differ by type of hazard (e.g. daytime temperatures or night-time minimum temperatures), exposure (e.g. exertional or classical heat stress), by type of vulnerability (e.g. age, sex, access to resources, chronic illness, and new reporting of symptoms) and design interventions accordingly. There is significant scope to improve existing responses. For instance, several existing heat-health warning systems rely on meteorological cut-offs only. Where sub-groups are differentially vulnerable, baseline health statues and activity levels need targeted attention.

Responses are typically developed in terms of disaster management, rather than taking on board long-term interventions. Long-term interventions sustainably reduce heat-health risks while often being the means to harness multiple co-benefits, such as advancing nature-based solutions, promoting green infrastructure, or reducing air pollution. Over the last one decade, several cities in northern India, ranked among the top 10 most polluting cities in the world, which potentially adds to the urban heat island effect; increased ozone and PM10 increase adverse respiratory outcomes. Empirical evidence on whether and how air pollution interacts with heat and, may magnify the health risks, needs strengthening in South Asia. Planned responses to ensure adequate sanitation and overcome water shortages during summer months can reduce gender-based vulnerabilities related to access, and for the workplace can prevent health and economic risks. Health care delivery systems in terms of hospital infrastructure, personnel, supply chains, patient support, and transportation have to be heat-resilient, to ensure functionality under extreme heat stress conditions.

## 5. CONCLUSION: SUMMARY AND OPPORTUNITIES FOR POLICY ACTION

### Summary

The paper contextualized the evidence available in India in terms of the determinants and key stressors for heat-health risks. These straddle physical, social, biological and institutional dimensions. Transitions in demography, urbanisation, and lifestyles can have consequences that lead to either positive or negative outcomes. Stressors interact amongst themselves, and also with ambient heat conditions to determine health outcomes. This emerging evidence, in combination with the early findings on exacerbation of chronic illnesses, has critical relevance for designing policy actions and, potentially, to modify current practices.

The results from the empirical analysis revealed that 20 of the 63 self-reported illnesses in the dataset (32%) were statistically significantly more likely to occur in summer than in the non-summer months. The findings are particularly significant because these are based on the NSS survey, which is amongst India’s best designed and executed large sample surveys, covering more than half a million surveyed individuals from across India. The differentials in reporting between summer and non-summer months are of critical concern, because these constitute major disease burdens for India, including heart diseases, hypertension, and diabetes. Further, the fact that occurrence is relatively high during summer months amongst older adults, females, and outdoor workers implies that multiple options are available for managing these risks and lowering disease burdens for the most vulnerable. The occurrence of hypertension increased from 74 to 96 per 1000, and diabetes from 73 to 90 per 1000 in the summer months amongst older adults, as compared with the non-summer months. The older population was also significantly vulnerable to heart diseases in summer months, while female populations and outdoor workers were additionally vulnerable to mental health related concerns. There is an opportunity for tailored policies for reducing disease burdens through effective public health planning and implementation, furthering the goals of national health policies [34] and helping achieve the Sustainable Development Goals.

There is enough available evidence to prompt meaningful action on reducing risks in both high- and low-income contexts. These include the health impacts of rising maximum and minimum temperatures, increasing frequency of heat waves, outdoor work exposure, access to cooling resources and healthcare. Some stressors are well studied in HICs but are not well understood in the context of LMICs. For example, the impact of biological vulnerabilities such as pre-existing Illnesses, sweating disorders, and medications, in relation to heat are well-established in the context of HICs but lacking in Indian literature. Some stressors such as rise in humidity and biological differences (sex) indicate divergences with no conclusive unidirectional findings, another area where research is needed to ensure timely actions and policy responses, aiming to strengthen systemic heat health preparedness.

Knowledge gaps are most prominent for LMICs, including India. Country specific studies are needed to address these knowledge gaps and establishing the divergent pathways by which heat stress turns into health risks. For instance, in many north-eastern and coastal regions in India, humidity is pronounced and more studies are needed on its role in localized conditions.

Similarly, the widespread exposure to high levels of air pollution in Indian cities intensifies biological vulnerability, creating compounded risks that require focussed research.

### Opportunities for policy action

#### Resilient health care systems

Surveillance and monitoring of heat-health outcomes, capacity-building and training of health care providers, and ensuring a heat resilient health care system can be challenging in LMICs, due to the inter-connected nature of heat stressors. An approach that integrates health actors into the broader framework of heat preparedness is crucial to build a healthcare system that can tackle HRIs and is resilient to heat health shocks. In most LMICs, within the public health system itself, there exist multiple vertical policies and programmes with hard targets that render the system inadequately prepared to tackle heat stress. Heat is an overarching concern cutting across dedicated programmes on communicable and non-communicable diseases. A strong inter-sectoral coordination mechanism would better enable the health system to deal with heat stress amongst vulnerable populations. Important learnings are available from HICs with health systems having evolved towards longer term resilience and sustained response mechanisms.

#### Integration of health actors

Integrating health sector actors in designing beneficial interventions outside the health sector could make planned responses more effective. Examples include awareness creation and adoption of local modifications to the built environment, such as green spaces and transportation changes that reduce risks by lowering air pollution and temperatures, thereby reducing cardiovascular risk. Improvements in socio-economic conditions of housing, neighborhoods, infrastructure, and access to care can be promoted with engagement of health sector actors in promoting principles of health equity.

#### Capacity Building and Awareness creation

Education and skill building to facilitate uptake of health adaptation activities that tends to be skewed towards younger adults and urban areas enables better protection from heat and lowered heat-health risks at both homes and work-places.

#### Integrating short and long term plans

Heat action plans and national level health adaptation policies are gaining relevance globally. In India, Heat Action Plans (HAP), have been the primary focus in terms of planned policy responses for addressing adverse heat-health outcomes. It is worrying that existing plans and practices failed to prevent heat-related deaths and complications in 2024. Responsive actions are more than a set of disaster response actions. As high heat events intensify and spread out over the country, a systemic visioning is required. Emergency response preparedness with plans that are to be operationalized during specific heatwave events lower excess mortality, and need to be complemented with long term, intersectoral plans and interventions that focus on reducing morbidity and mortality. Examples include policies for improving working conditions for outdoor labor and school regulations, urban design, building and settlement plans, and ensuring energy security for emergency health care services. Effectively addressing the diverse range of climate related health risks requires plans and policies at the nexus of health and inter-related sectors to be well-synchronized.

#### Decentralized Governance, with Data and Surveillance

Heat-health management requires data analysis and innovation based on surveillance to better understand its distribution across geographies, economic, and social determinants, alongside availability of health facilities that align with health-seeking behaviour. The WHO identifies the health authority as the lead agency for HRI management. The authority should be able to draw upon existing systems and arrangements for emergency preparedness and response, work in an intersectoral manner with appropriate coordination arrangements while ensuring that all actors have enough information and resources to undertake the respective sectoral activities. The well-recognized strengths of local health authorities include direct accountability to local communities, coordination with other social care and basic services and ability to respond to health inequities; they will need to be strengthened with health system competences, appropriate resources and flexibility in decision making. For instance, India’s rapidly expanding digital and weather forecasting capabilities can be used to shape an information network for timely dissemination of alerts, in local languages and for use by local actors.

#### Communication

A key weakness that needs to be urgently addressed is a two-way heat risk communications capability. Top-down communication addresses broad heat-health risks, weather, roles and responsibilities for the implementation actors and behavioral guidance for at-risk groups. On the other hand, bottom-up communication includes event reporting, survey of vulnerable groups, and availability of frontline resources such as cooling centers. Segmentation of audience and customized communications can be based on age, workplace, those with known chronic diseases, and neighborhoods with specific locational disadvantages. High-quality data on socioeconomic status, infrastructure such as power and water and housing, facilitate customized communications. Effective communications require an understanding of the “risk signature” (of heat-health) among vulnerable groups, namely, the way individuals and communities’ reason practically about specific risks. Subjective perceptions of risk are shaped by a complex interplay of multiple factors: media discourse, lived experience, risk aversion patterns, ethnicity (including migration experiences), and familiarity (how frequently one is exposed to a hazard). Vulnerable groups should be prioritized for outreach at the beginning of summer, communicated the heat-health risks, provided guidance on coping and specific information on availability and location of specific health and social services. Protecting specific occupational groups entail a mix of strategies – working with their employers and trade unions, facilitating implementation of relevant regulatory measures.

#### Strengthening urban health system resilience

Urban populations are at greater risk of heat-health consequences than rural ones. India’s urban health system is plagued by a plethora of challenges – multiplicity of agencies, high degree of privatization, varied strengths of municipal bodies, and a still-evolving National Urban Health Mission. Even as Indian states begin developing state action plans on climate change and health, responding to the heat-health challenges provides an opportunity to strengthen resilience of health systems and promote health outcomes. Expanding the evidence and understanding on how heat impacts human health is essential for risk management. New approaches such as health in all policies, greening the health and pharmaceutical sectors, housing and urban planning to reduce urban heat island effects, can draw upon this evidence for cost-effective adaptation and mitigation actions that deliver on health goals and universal health coverage.

## Ethics Statement

The study is approved by the Institutional Ethics Committee of the Institute of Economic Growth, certification dated Aug 27, 2024; Grant Number UK [226740/Z/22/Z]. This study is based on publicly available secondary data sources, and no patients were involved.

## Contributors

PD, RD, and KE were responsible for conceptualization and design. All authors contributed to methods, data collection, data analysis and interpretation of the results. All authors contributed to the initial writing while PD, RD, KE, GS and SP were also involved in critically reviewing and revising the article. All authors approve the final version and are accountable for the same.

## Acknowledgement

The authors thank Khushboo Aggarwal and Rupam Tiwari for their help with some parts of the data collation.

## Data Availability Statement

All data used are available in a public, open access repository. Data for the National Sample Survey 75^th^ health round is publicly available online at https://microdata.gov.in/NADA/index.php/catalog/152/get-microdata. District level temperature data used in the study is also publicly available online (http://data.icrisat.org/dld/src/environment.html).

