## Supplemental Material for "Harms from Heat-Health Risks: Morbidity Evidence from India and Global Learnings for Policy Action"

## SM1

### Summary of key studies on heat related mortality in India

#### Table 1| Mortality Literature review

| Author | Year of study | Area of study/Population group | Summary |
|---|---|---|---|
| Burgess et al 2014 | 1957 to 2000 | India (district level) | Burgess et al utilize district-level daily weather (IMD) and annual mortality data for 334 districts covering the whole of India for 1957 to 2000. The study reveals a stark inequality: hot days substantially increase mortality in rural, but not urban, India. This vulnerability in rural areas is attributed to the impact of hot days during the growing season, which reduce agricultural productivity and wages. |
| Desai et al 2015 | 2001–2012 | Urban ares of Surat, a coastal city in India | The study analyzed 36,167 deaths over 12 summers (2001–2012), revealing a significant correlation between all-cause mortality and ambient heat. Notably, mortality increased by 11% when temperatures exceeded 40°C, and by 18% on high-risk heat days (extreme danger), with the Heat Index (HI) being a more critical predictor than maximum temperature. The study emphasizes the urgent need for adaptation measures to mitigate heat-related health impacts in vulnerable urban populations. |
| Bont et al 2024 | 2008-2019 | Ten cities in India: Ahmedabad, Bangalore, Chennai, Delhi, Hyderabad, Kolkata, Mumbai, Pune, Shimla, and Varanasi | Among ~ 3.6 million deaths, the study observed that temperatures above 97th percentile for 2-consecutive days was associated with a 14.7 % (95 %CI, 10.3; 19.3) increase in daily mortality. Shorter and less |

| | | | |
|---|---|---|---|
| | | | intense definitions of heatwaves resulted in a higher estimated burden of heatwave-related deaths. |
| Rathi & Sodani 2021 | 2006 to 2015 | Hyderabad city population. | The study uses data on temperature and all-cause mortality for at least ten years for summer months and found an increase of 16% and 17% per day mean all-cause mortality at the maximum temperature of ≥40°C and for extreme danger days (Heat Index >54°C) respectively. |
| Nori et al 2019 | 2000-2012 | Northwest India | Using daily all-cause mortality for 4 communities in Northwest India, the study found reveals that total mortality increased 18.1% during heat wave days compared with non-heat-wave days |
| Azhar et al 2014 | May 2010 | Ahmedabad, Gujarat, India | An analysis of the May 2010 heatwave in Ahmedabad, Gujarat, revealed a substantial 43.1% increase in all-cause mortality, with 1,344 excess deaths. This study found strong correlations between daily maximum temperature and mortality during the hottest months (April, May, June), indicating a significant impact of extreme heat on mortality even in this specific city. |

**SM2**

**Data Source 1| District-wise monthly maximum and minimum temperatures**

The temperature data used in this study is sourced from the International Crops Research Institute for the Semi-Arid Tropics (ICRISAT), a global research organization focused on improving livelihoods in dryland tropics. From its extensive data repositories, ICRISAT provides high-resolution weather data, including maximum and minimum temperatures, rainfall, and other climatic indicators, alongside comprehensive socioeconomic and crop data, detailing production, irrigation, soil types, and market access, across 571 districts in India. Available monthly highest maximum and lowest minimum temperature data was extracted for districts across India, spanning July 2017 to June 2018.

**Data Source Link:** http://data.icrisat.org/dld/src/environment.html

**Figure 1| Distribution of 561 Indian Districts by Monthly 'Lowest Minimum Temperature (LMIN)' Bins**

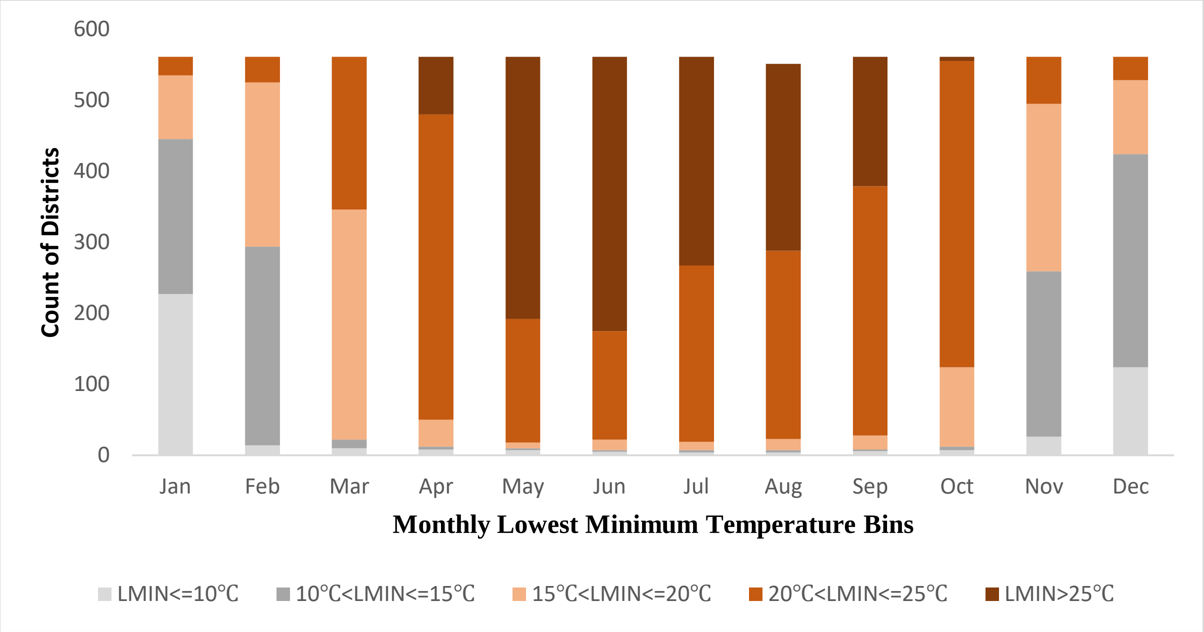

**Table 1| Independent Samples T-Test for Monthly Highest Maximum Temperature (Summer vs. Non-Summer Months)**

| **Group** | **Observations** | **Mean** | **Standard Error** | **Standard Deviation** | **95% Confidence interval** | |
|---|---|---|---|---|---|---|
| Non-Summer | 4488 | 29.15461 | 0.064 | 4.347 | 29.02737 | 29.28185 |
| Summer | 2244 | 35.45419 | 0.102 | 4.835 | 35.25401 | 35.65437 |
| Combined | 6,732 | 31.25447 | 0.065876 | 5.405008 | 31.12533 | 31.38361 |
| diff | | -6.29958 | 0.120965 | | -6.53673 | 6.06242 |
| diff = mean(Non-Summ) – mean (Summer M) t = -52.0777<br>Ho: diff = 0 Satterthwaite's degrees of freedom = 4088.88 | | | | | | |
| **Ha: diff < 0**<br>**Pr(T < t) = 0.0000** | **Ha: diff != 0**<br>**Pr(\|T\| > \|t\|) = 0.0000** | | **Ha: diff > 0**<br>**Pr(T > t) = 1.0000** | | | |

Table 1 presents the results of an independent samples t-test comparing the mean highest maximum temperatures of the month between summer and non-summer periods. A statistically significant difference was observed, indicating that summer months consistently exhibit different maximum temperatures than non-summer months.

**SM3**

The available evidence on the factors determining heat-related health risks was categorized as well-established, partially established, or not established based on a 2 stage review process. At the first stage, availability of evidence from at least one LMIC country, apart from India, was considered as an essential criterion. If this criterion was unfulfilled, the global evidence on the corresponding risk factor was classified as not established. At the second stage, the strength of evidence on the risk factor was additionally considered. Table 1 presents the classification. Where evidence was available for more than 2 LMICs as well as more than 2 HICs, and the studies had similar findings, these were classified as well-established. If there were 1 or 2 studies from LMICs other than India, the evidence was classified as partially established, subject to there being at least 2 HIC studies, and similarity in findings across all the studies. Where there were dissimilarities in the findings in terms of the directionality of the impacts of heat on human health irrespective of an LMIC or HIC context, the evidence for the corresponding risk factor was considered as non-established. This held even if a single study reported a contrary or ambiguous finding on the concerned risk factor. Similarly, for India, availability of more than 4 studies with similar findings was considered to be well-established, while having 4 to 2 studies with similar findings was partial evidence, and instances of dissimilar findings or having less than 2 studies was considered as evidence not established.

**SM4**

**Supporting tables and figures for Empirical analysis**

**Figure 1| Differentials in reported illnesses during summer and non-summer months, All Individuals**

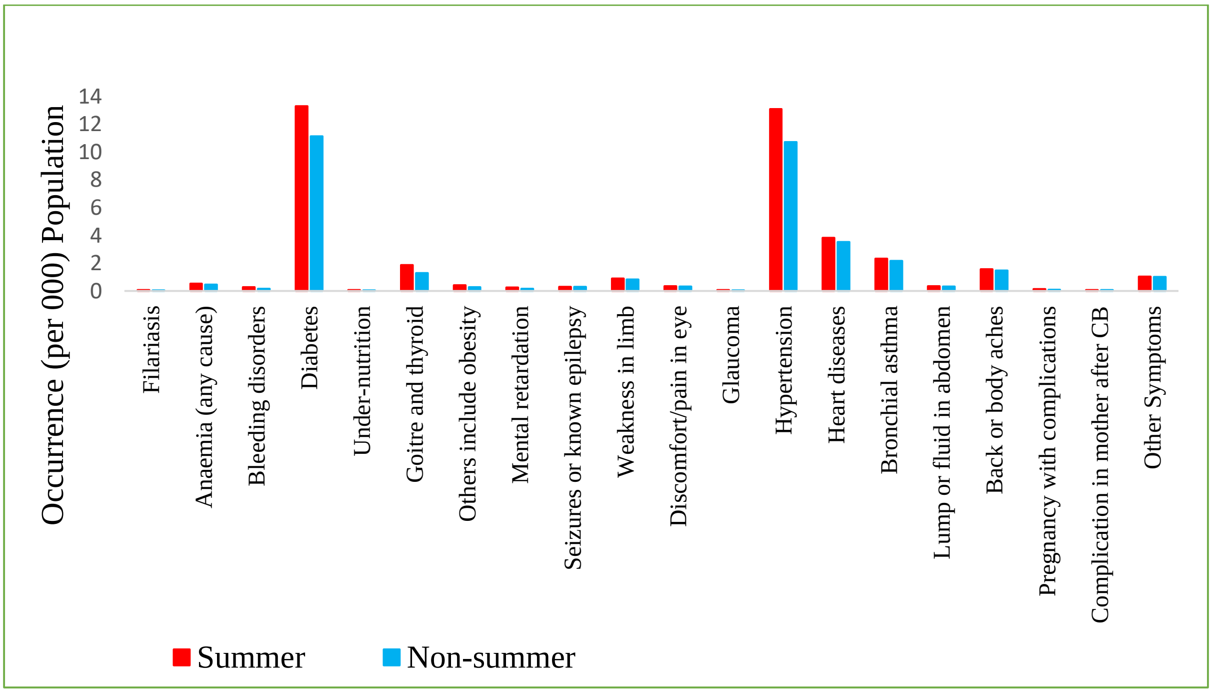

**Table 1| Demographic details from the NSS survey data**

| Variable | Description | Sample size | Percentage share |
|---|---|---|---|
| Age (years) | 0-5 | 70,258 | 12.7 |
| | 6-14 | 85,389 | 15.4 |
| | 15-24 | 94,676 | 17.1 |
| | 25-45 | 1,85,388 | 33.3 |
| | 46-59 | 76,879 | 13.8 |
| | 60 and above | 42,762 | 7.7 |
| Gender | Male | 2,83,200 | 50.99 |
| | Female | 2,72,115 | 49.00 |
| | Transgender | 37 | 0.01 |
| Occupation | Indoor | 3,12,771 | 56.3 |
| | Outdoor | 2,42,581 | 43.7 |

**Table 2| chi-square test for illnesses, All India, all individuals**

| **Illness** | **Groups** | **$\chi 2$ Value** | ***p*-Value** |
|---|---|---|---|
| **Hypertension** | Summer | 60.0098 | 0.000** |
| | Non-summer | | |
| **Heart disease** | Summer | 3.1607 | 0.075* |
| | Non-summer | | |
| Filariasis | Summer | 0.2165 | 0.642 |
| | Non-summer | | |
| Anaemia | Summer | 0.5724 | 0.449 |
| | Non-summer | | |
| **Bleeding disorders** | Summer | 6.5155 | 0.011** |
| | Non-summer | | |
| **Diabetes** | Summer | 48.4097 | 0.000** |
| | Non-summer | | |
| Under-nutrition | Summer | 1.9487 | 0.163 |
| | Non-summer | | |
| **Goitre and thyroid** | Summer | 27.7368 | 0.000** |
| | Non-summer | | |
| **Others including Obesity** | Summer | 6.2960 | 0.012** |
| | Non-summer | | |
| **Mental Retardation** | Summer | 4.2898 | 0.038** |
| | Non-summer | | |
| Seizures or known epilepsy | Summer | 0.0162 | 0.899 |
| | Non-summer | | |
| Weakness in limb/Immobility | Summer | 0.5270 | 0.468 |
| | Non-summer | | |

| | | | |
|---|---|---|---|
| Discomfort/pain in eye | Summer | 0.3289 | 0.566 |
| | Non-summer | | |
| Glaucoma | Summer | 2.6039 | 0.107 |
| | Non-summer | | |
| Bronchial asthma | Summer | 1.2647 | 0.261 |
| | Non-summer | | |
| Lump or fluid in abdomen | Summer | 0.2692 | 0.604 |
| | Non-summer | | |
| Back or body aches | Summer | 0.5305 | 0.466 |
| | Non-summer | | |
| Pregnancy complications | Summer | 1.0197 | 0.313 |
| | Non-summer | | |
| Complications in mother after child birth | Summer | 0.8003 | 0.371 |
| | Non-summer | | |
| Any Other Symptom | Summer | 0.0162 | 0.899 |
| | Non-summer | | |

Source: Author's Calculation ** denotes significance at 5% alpha level ($p$<0.05), * denotes significance at 10% alpha level ($p$<0.10)

**Table 3| Odds Ratio for seven illnesses prevalent in summer season**

| **Illness** | **Groups** | **Odds Ratio** | ***p*-Value** |
|---|---|---|---|
| **Hypertension** | Summer | 1.288 | 0.000** |
| | Non-summer | | |
| **Heart disease** | Summer | 1.094 | 0.062* |
| | Non-summer | | |
| **Bleeding disorders** | Summer | 1.596 | 0.010** |
| | Non-summer | | |
| **Diabetes** | Summer | 1.231 | 0.000** |
| | Non-summer | | |
| **Goitre and thyroid** | Summer | 1.457 | 0.000** |
| | Non-summer | | |
| **Others including Obesity** | Summer | 1.443 | 0.014** |
| | Non-summer | | |
| **Mental Retardation** | Summer | 1.483 | 0.038** |
| | Non-summer | | |

Source: Author's Calculation ** denotes significance at 5% alpha level ($p$<0.05), * denotes significance at 10% alpha level ($p$<0.10)

**Table 4| Summary information on number of respondents reporting illness in the 15 days prior to the survey date**

| Illness | Older adults (60 years and above) | | Females | | Outdoor Workers | |
|---|---|---|---|---|---|---|
| | Summer (12,874) | Non-summer (29,888) | Summer (85,792) | Non-summer (1,86,323) | Summer (76,209) | Non-summer (166,372) |
| Hypertension (6,369) | 1,231 | 2,206 | 1,273 | 2,230 | 420 | 802 |
| Diabetes (6,563) | 1,165 | 2,179 | 1,142 | 2,102 | 397 | 749 |

Notes: Figures in brackets indicate the number of individuals in the respective category

**SM5**

**Table 1| Supporting Table on heat health determinants**

| **Stressors** | **Factors Influencing Adverse Outcomes** | **Source (HIC)** | **Source (LMIC)** | **India** |
|---|---|---|---|---|
| **Heat Hazard (Direct)** | **Increasing Maximum Temperatures** | O'Neill & Ebi 2009; Cheng et al. 2014; Deschênes & Greenstone 2011; Gasparrini et al. 2012 | McElroy et al. 2022; Lakhani et al. 2024; Siddik, & Rahman, 2014 | Rathi et al .2021; de Bont et al 2024; Burgess et al 2014; Azhar et al 2014; Jaswal et al 2017 |
| | **Increasing Minimum Temperatures** | Cheng et al 2014 *[Meta Analysis]* | Cheng et al 2014 *[Meta Analysis];* Siddik & Rahman 2014 | Burgess et al 2014; Mall et al 2021 |
| | **Rise in Humidity** | Armstrong et al. 2019; Baldwin et al. 2023; Lepeule 2018 | McMahon et al | Fritz 2025 |
| | **Rise in Heatwave Occurrence** | Habeeb et al 2015; Russo et al 2015; Lemonsu et al 2014 | Green et al 2019; Sapari et al 2023; Haque et al 2024 | de Bont et al 2024; Maurya et al 2024; Singh et al 2021; Saunik & Shaw; Thakkar et al 2025 |
| **Exposure** | **Outdoor work** | Ioannou et al 2022 [Meta Analysis]; Habibi et al 2024 | Ioannou, L. G. et al 2022 *[Meta Analysis];* Habibi et al 2024 | Ioannou et al 2022 [Meta Analysis]; Habibi et al 2024; Sahu S et al 2013; Patel H et al 2006; Khetan et al. 2024 |

| | | | | |
|---|---|---|---|---|
| | **Lack of ventilation** | Wargocki, P. (2013).; Lenzer, B. et al 2020; Jensen, C. A. et al 2017; Lundgren Kownacki et al 2019 | Samuelson et al 2020 [SIMULATION BASED] | Samuel et al 2017; Manu et al 2019; Srikonda & Dokiparty (2010, September) |
| | **Indoor heat load** | Tham et al 2020; Anderson et al 2013; Basu 2009; McMichael et al 2008 | Tham et al 2020; Teare et al 2020; McMichael,et al 2008; Samuelsonet al 2000 | Mukhopadhyay et al 2021; McMichael et al 2008; Samuelson et al 2020 *[SIMULATION BASED];* Samuel et al 2017; Manu et al 2019 |
| **Biological/Physiological Vulnerability** | **Age** | Basu 2009; Basu et al 2015; Bell et al 2008; Lundgren et al 2019; Baccini et al 2008 | Basu 2009; Bell et al 2008; Gouveia et al 2003; Yu et al 2012 | Weitz et al 2022; Khetan et al 2024; Agarwala et al 2020; Lala, B., & Hagishima, A. (2023). |
| | **Pre-existing Illnesses (Chronic diseases (e.g., cardiovascular, respiratory, kidney conditions, psychiatric conditions) are at higher risk of worsening.)** | Lõhmus et alKim et al 2014; Woodland et al 2023; Kaiser et al 2001 | Hall & Hall 2021 *[Physiological Evidence]* | Shrikhande et al 2022 |
| | **Disorders of Sweating** | Kovats, R. S., & Hajat, S. (2008).; Hall, J. E. (Ed.). (2021) | Hall & Hall (2021). *[Physiological Evidence]* | |
| | **Medication** | Layton et al 2020; Hermesh et al 2000; Kovats, R. S., & Hajat, S. (2008); Kaiser et al 2001; Westaway et al 2015 *[physiological evidence]* | Hospers, L. et al 2024 | |
| | **Onset of New Illnesses (CVD, Mental Illness)** | Mora, C et al 2017; Campbell et al 2018; Morris, A. & Patel, G (2023). *[physiological evidence]* | Campbell et al 2018; Green et al 2019; Razzak et al 2022 | Khetan et al 2024; Venugopal et al 2016; Shrikhande et al 2022; Mukhopadhyay et al 2022; Varghese et al 2005 |
| **Socio-economic Vulnerability** | **Income** | Chakraborty et al 2019[meta analysis] | Chakraborty et al 2019 | Venugopal et al 2016; Nanda et al 2022 Das, S. (2015); Tran et al 2013; Somanathan et al 2021 |
| | **Housing Quality** | Howden-Chapman et al 2023; Samuelson et al 2020 [simulation based] ; Lawrence, R. J. (2004). | Kajjoba et al 2022 | Khetan et al 2024; Samuel et al 2017; Pradyumna et al 2018 |

| | | | | |
|---|---|---|---|---|
| | **Access to Cooling Resources (Energy, Equipment, Water)** | Song et al 2024; Bedi et al 2022; Widerynski et al 2017 | Tawsif et al 2022; Hasan et al 2021 [Scoping Review] | Khetan et al 2024; Venugopal et al 2016; Mukhopadhyay & Weitz (2022); Pradyumna et al 2018 |
| | **Access to healthcare** | Mullins & White (2020); Lee et al 2024; Woolf et al 2023 | Cheng, & Sha (2024); Sapari et al 2023 [Systemetic Review] | Pradyumna et al 2018; Dureja & Jain (2025); Nanda et al 2022; Tran et al 2013; Roy et al 2017 |
| | **Neighbourhood (Green and blue spaces)** | Targino et al 2019; Ebi, K. L., & Bowen, K. (2023); Kumar et al 2024 | Pritipadmaja et al 2023; Kumar, & Shekhar (2021); Hunter et al 2023; Hosen et al 2025 | Krishnaswamy et al 2025; Pritipadmaja et al 2023; Khetan et al 2024 |
| **Sex and Gender Vulnerability** | **Biological Differences** | Hall, J. E. (2021); Druyan et al 2012; Lakhoo et al 2025 | Bhan et al 2022; Lakhoo et al 2025; Bell et al 2008 | Venugopal et al 2016; Venugopal et al 2016; Choudhary RK et al; Kumar A, Singh DP. |
| | **Sociocultural Factors** | Seebass, K. (2017); Florido Ngu et al 2021; Lundgren Kownacki et al 2019; Brown, S., & Walker, G. (2013); Madrigano et al 2015 | Zhu et al 2023 | Mitchell et al 2021; Venugopal et al 2016; Khetan et al 2024; Venugopal et al 2016 |
| | **Access to Resources** | Kadio et al 2024; Howells et al 2025; Johar et al 2025 | Lusambili, A. et al 2024; Bethancourt et al 2021; Sampson et al 2013 | Khetan et al 2024; Mukhopadhyay et al 2022; Trahan et al 2023; Dasgupta et al 2024; Venugopal et al 2019; Nanda et al 2022 |
| **Responses** | **Heat Health Warning systems** | Matzarakis et al 2019; Ebi (2007); Lowe et al 2011; Rao et al 2025 | RIMES. (2024) | Golechha et al 2021; Trahan 2023 |
| | **Cooling Interventions** | Jay et al 2021; Dearman et al 2024; Coutts et al 2013; Limaye, V. S. (2023). | Bunker et al 2024; Montaldo et al 2015; Shahrujjaman et al 2025 | Pradyumna et al 2018; Debnath et al 2021; Vellingiri et al 2020; Chandrashekhar, V. (2023); Arumugam et al 2014 |
| | **Public Awareness** | Johar et al 2025 [Systematic Review]Razzak et al 2022 | Johar et al 2025[Systematic Review] | Das & Smith, (2012); Maheshwari (2022); Das (2016). ; Venugopal et al 2016; Pradyumna et al 2018 |
| | **Policy and Regulations** | Bolitho, A., & Miller, F. (2017); Hatvani-Kovacs et al 2018; Hasan et al 2021; Tomlinson et al 2011 | Hasan et al 2021 [Three LMICs]; Quinn et al 2022 | Pradyumna et al 2018; Singh et al 2024 ; Golechha et al 2024 ;Directorate of Environment and Climate Change (DoECC), State Action Plan on Climate Change. |

| | | | | |
|---|---|---|---|---|
| | **Air Pollution (Synergistic effects)** | Brian 2005 | | |
| | **Reducing Urban Heat Island Effect** | Tomlinson et al 2011, Targino et al 2019, Ebi & Bowen 2023, Kumar et al 2024 | Jahan, I. (2024); Hanif et al 2022 | Dwivedi, A., & Mohan, B. K. (2018); Khare et al 2021 |

### Supporting References

#### High Income Countries (HIC)

O'Neill MS, Ebi KL. Temperature extremes and health: impacts of climate variability and change in the United States. J Occup Environ Med. 2009;51(1):13-25.

Cheng J, Xu Z, Zhu R, Wang X, Jin L, Song J, Su H. Impact of diurnal temperature range on human health: a systematic review. Int J Biometeorol. 2014;58:2011-24.

Deschênes O, Greenstone M. Climate change, mortality, and adaptation: Evidence from annual fluctuations in weather in the US. Am Econ J Appl Econ. 2011;3(4):152-85.

Gasparrini A, Armstrong B, Kovats S, Wilkinson P. The effect of high temperatures on cause-specific mortality in England and Wales. Occup Environ Med. 2012;69(1):56-61.

Cheng J, Xu Z, Zhu R, Wang X, Jin L, Song J, Su H. Impact of diurnal temperature range on human health: a systematic review. Int J Biometeorol. 2014;58:2011-24. [Meta Analysis]

Armstrong B, et al. The role of humidity in associations of high temperature with mortality: A multicountry, multicity study. Environ Health Perspect. 2019;127:097007-1-097007–8.

Baldwin JW, et al. Humidity's Role in Heat-Related Health Outcomes: A Heated Debate. Environ Health Perspect. 2023;131.

Lepeule J, Litonjua AA, Gasparrini A, Koutrakis P, Sparrow D, Vokonas PS, Schwartz J. Lung function association with outdoor temperature and relative humidity and its interaction with air pollution in the elderly. Environ Res. 2018;165:110-7.

Habeeb D, Vargo J, Stone B. Rising heat wave trends in large US cities. Nat Hazards. 2015;76:1651-65.

Russo S, Sillmann J, Fischer EM. Top ten European heatwaves since 1950 and their occurrence in the coming decades. Environ Res Lett. 2015;10(12):124003.

Lemonsu A, Beaulant AL, Somot S, Masson V. Evolution of heat wave occurrence over the Paris basin (France) in the 21st century. Clim Res. 2014;61(1):75-91.

Ioannou LG, Foster J, Morris NB, Piil JF, Havenith G, Mekjavic IB, et al. Occupational heat strain in outdoor workers: a comprehensive review and meta-analysis. Temperature. 2022;9(1):67-102. [Meta Analysis]

Habibi P, Razmjouei J, Moradi A, Mahdavi F, Fallah-Aliabadi S, Heydari A. Climate change and heat stress resilient outdoor workers: findings from systematic literature review. BMC Public Health. 2024;24(1):1711.

Wargocki P. The effects of ventilation in homes on health. Int J Ventil. 2013;12(2):101-18.

Lenzer B, Rupprecht M, Hoffmann C, Hoffmann P, Liebers U. Health effects of heating, ventilation and air conditioning on hospital patients: a scoping review. BMC Public Health. 2020;20(1):1287.

Jensen CA, Cadorel X, Chu A. Ventilation for reduced heat stress in apartments. Back to the Future: The Next. 2017;50:615-24.

Lundgren Kownacki K, Gao C, Kuklane K, Wierzbicka A. Heat stress in indoor environments of scandinavian urban areas: A literature review. Int J Environ Res Public Health. 2019;16(4):560.

Tham S, Thompson R, Landeg O, Murray KA, Waite T. Indoor temperature and health: a global systematic review. Public Health. 2020;179:9-17.

Anderson M, Carmichael C, Murray V, Dengel A, Swainson M. Defining indoor heat thresholds for health in the UK. Perspect Public Health. 2013;133(3):158-64.

Basu R. High ambient temperature and mortality: A review of epidemiologic studies from 2001 to 2008. Environ Health. 2009;8(40). doi:10.1186/1476-069X-8-40.

McMichael AJ, Wilkinson P, Kovats RS, Pattenden S, Hajat S, Armstrong B, et al. International study of temperature, heat and urban mortality: the ‘ISOTHURM’project. Int J Epidemiol. 2008;37(5):1121-31.

Basu R, Dominici F, Samet JM. Temperature and mortality among the elderly in the United States: a comparison of epidemiologic methods. Epidemiology (Camb Mass). 2005;16(1):58–66. doi:10.1097/01.ede.0000147117.88386.fe.

Bell ML, O’neill MS, Ranjit N, Borja-Aburto VH, Cifuentes LA, Gouveia NC. Vulnerability to heat-related mortality in Latin America: a case-crossover study in Sao Paulo, Brazil, Santiago, Chile and Mexico City, Mexico. Int J Epidemiol. 2008;37(4):796-804.

Baccini M, Biggeri A, Accetta G, Kosatsky T, Katsouyanni K, Analitis A, Anderson HR, Bisanti L, D'Ippoliti D, Danova J, Forsberg B, Medina S, Paldy A, Rabczenko D, Schindler C, Michelozzi P. Heat effects on mortality in 15 European cities. Epidemiology. 2008;19(5):711–9. doi:10.1097/EDE.0b013e318176bfcd.

Lõhmus M. Possible biological mechanisms linking mental health and heat—a contemplative review. Int J Environ Res Public Health. 2018;15. Preprint at: https://doi.org/10.3390/ijerph15071515.

Kim SH, Jo SN, Myung HN, Jang JY. The effect of pre-existing medical conditions on heat stroke during hot weather in South Korea. Environ Res. 2014;133:246-52.

Woodland L, Ratwatte P, Phalkey R, Gillingham EL. Investigating the health impacts of climate change among people with pre-existing mental health problems: a scoping review. Int J Environ Res Public Health. 2023;20(8):5563.

Kaiser R, Rubin CH, Henderson AK, Wolfe MI, Kieszak S, Parrott CL, Adcock M. Heat-related death and mental illness during the 1999 Cincinnati heat wave. Am J Forensic Med Pathol. 2001;22(3):303-7.

Kovats RS, Hajat S. Heat stress and public health: a critical review. Annu Rev Public Health. 2008;29:41-55.

Hall JE, editor. Guyton & Hall. Tratado de fisiología médica. Elsevier Health Sciences; 2021.

Layton JB, Li W, Yuan J, Gilman JP, Horton DB, Setoguchi S. Heatwaves, medications, and heat-related hospitalization in older Medicare beneficiaries with chronic conditions. PLoS One. 2020;15(12):e0243665.

Hermesh H, Shiloh R, Epstein Y, Manaim H, Weizman A, Munitz H. Heat intolerance in patients with chronic schizophrenia maintained with antipsychotic drugs. Am J Psychiatry. 2000;157(8):1327-9.

Westaway K, Frank O, Husband A, McClure A, Shute R, Edwards S, et al. Medicines can affect thermoregulation and accentuate the risk of dehydration and heat-related illness during hot weather. J Clin Pharm Ther. 2015;40(4). [PHYSIOLOGICAL EVIDENCE]

Mora C, Counsell CWW, Bielecki CR, Louis LV. Twenty-seven ways a heat wave can kill you: Deadly heat in the era of climate change. Circ Cardiovasc Qual Outcomes. 2017;10.

Campbell S, Remenyi TA, White CJ, Johnston FH. Heatwave and health impact research: A global review. Health Place. 2018;53:210-8.

Morris A, Patel G. Heat Stroke. In: StatPearls. Treasure Island (FL): StatPearls Publishing; 2023. [PHYSIOLOGICAL EVIDENCE]

Chakraborty T, Hsu A, Manya D, Sheriff G. Disproportionately higher exposure to urban heat in lower-income neighborhoods: a multi-city perspective. Environ Res Lett. 2019;14(10):105003. [META ANALYSIS]

Howden-Chapman P, Bennett J, Edwards R, Jacobs D, Nathan K, Ormandy D. Review of the impact of housing quality on inequalities in health and well-being. Annu Rev Public Health. 2023;44(1):233-54.

Samuelson H, Baniassadi A, Lin A, González PI, Brawley T, Narula T. Housing as a critical determinant of heat vulnerability and health. Sci Total Environ. 2020;720:137296. [SIMULATION BASED]

Lawrence RJ. Housing and health: from interdisciplinary principles to transdisciplinary research and practice. Futures. 2004;36(4):487-502.

Song W, Ding Q, Huang M, Xie X, Li X. Meta-analysis study on the effects of personal cooling strategies in reducing human heat stress: Possible application to medical workers. J Build Eng. 2024;85:108685.

Bedi NS, Adams QH, Hess JJ, Wellenius GA. The role of cooling centers in protecting vulnerable individuals from extreme heat. Epidemiology. 2022;33(5):611-5.

Widerynski S, Schramm PJ, Conlon KC, Noe RS, Grossman E, Hawkins M, et al. Use of cooling centers to prevent heat-related illness: summary of evidence and strategies for implementation.

Mullins JT, White C. Can access to health care mitigate the effects of temperature on mortality?. J Public Econ. 2020;191:104259.

Lee J, Min J, Lee W, Sun K, Cha WC, Park C, et al. Timely accessibility to healthcare resources and heatwave-related mortality in 7 major cities of South Korea: a two-stage approach with principal component analysis. Lancet Reg Health West Pac. 2024;45.

Woolf S, Morina J, French E. The health care costs of extreme heat. 2023.

Targino AC, Coraiola GC, Krecl P. Green or blue spaces? Assessment of the effectiveness and costs to mitigate the urban heat island in a Latin American city. Theor Appl Climatol. 2019;136(3):971-984.

Ebi KL, Bowen K. Green and blue spaces: crucial for healthy, sustainable urban futures. Lancet. 2023;401(10376):529-530.

Kumar P, Debele SE, Khalili S, Halios CH, Sahani J, Aghamohammadi N, et al. Urban heat mitigation by green and blue infrastructure: Drivers, effectiveness, and future needs. Innov. 2024;5(2).

Druyan A, Makranz C, Moran D, Yanovich R, Epstein Y, Heled Y. Heat tolerance in women—reconsidering the criteria. Aviat Space Environ Med. 2012;83(1):58-60.

Lakhoo DP, Brink N, Radebe L, Craig MH, Pham MD, Haghighi MM, et al. A systematic review and meta-analysis of heat exposure impacts on maternal, fetal and neonatal health. Nat Med. 2025;31(2):684-94.

Seebass K. Who Is Feeling the Heat?: Vulnerabilities and Exposures to Heat Stress—Individual, Social, and Housing Explanations. Nat Cult. 2017;12(2):137-61.

Florido Ngu F, Kelman I, Chambers J, Ayeb-Karlsson S. Correlating heatwaves and relative humidity with suicide (fatal intentional self-harm). Sci Rep. 2021;11(1):22175.

Brown S, Walker G. Understanding heat wave vulnerability in nursing and residential homes. In: Comfort in a Lower Carbon Society. Routledge; 2013. p. 59-68.

Madrigano J, Ito K, Johnson S, Kinney PL, Matte T. A case-only study of vulnerability to heat wave–related mortality in New York City (2000–2011). Environ Health Perspect. 2015;123(7):672-8.

Kadio K, Filippi V, Congo M, Scorgie F, Roos N, Lusambili A, et al. Extreme heat, pregnancy and women's well-being in Burkina Faso: an ethnographical study. BMJ Glob Health. 2024;8(Suppl 3).

Howells M, Palmquist AE, Josefson C, Dancause K, Quinn E, Daniels L, Blair AFO. Climate change, evolution, and reproductive health: The impact of water insecurity and heat stress on pregnancy and lactation. Evol Med Public Health. 2025;eoaf008.

Johar H, Abdulsalam FI, Guo Y, Baernighausen T, Jahan NK, Watterson J, et al. Community-based heat adaptation interventions for improving heat literacy, behaviours, and health outcomes: a systematic review. Lancet Planet Health. 2025.

Matzarakis A, Laschewski G, Muthers S. The heat health warning system in Germany—Application and warnings for 2005 to 2019. Atmosphere. 2020;11(2):170.

Ebi KL. Towards an early warning system for heat events. J Risk Res. 2007;10(5):729-44.

Lowe D, Ebi KL, Forsberg B. Heatwave early warning systems and adaptation advice to reduce human health consequences of heatwaves. Int J Environ Res Public Health. 2011;8(12):4623-48.

Rao S, Chaudhary P, Budin-Ljøsne I, Sitoula S, Aunan K, Chersich M, et al. Evaluating the socioeconomic benefits of heat-health warning systems. Eur J Public Health. 2025;ckae203.

Jay O, Capon A, Berry P, Broderick C, de Dear R, Havenith G, et al. Reducing the health effects of hot weather and heat extremes: from personal cooling strategies to green cities. Lancet. 2021;398(10301):709-24.

Dearman C, Adams N, Sousa A, Pearce-Smith N, Petrokofsky C. Public health effectiveness of cooling centres in adverse hot weather events: a systematic review. Eur J Public Health. 2024;34(Suppl 3):ckae144-1367.

Coutts AM, Daly E, Beringer J, Tapper NJ. Assessing practical measures to reduce urban heat: Green and cool roofs. Build Environ. 2013;70:266-76.

Limaye VS. The hidden health costs of climate change: Accounting for extreme heat harms to women in the global South. PLoS Clim. 2023;2(8):e0000267.

Johar H, Abdulsalam FI, Guo Y, Baernighausen T, Jahan NK, Watterson J, et al. Community-based heat adaptation interventions for improving heat literacy, behaviours, and health outcomes: a systematic review. Lancet Planet Health. 2025. [Systematic Review]

Razzak JA, Agrawal P, Chand Z, Quraishy S, Ghaffar A, Hyder AA. Impact of community education on heat-related health outcomes and heat literacy among low-income communities in Karachi, Pakistan: a randomised controlled trial. BMJ Glob Health. 2022;7(1):e006845.

Bolitho A, Miller F. Heat as emergency, heat as chronic stress: policy and institutional responses to vulnerability to extreme heat. Local Environ. 2017;22(6):682-98.

Hatvani-Kovacs G, Bush J, Sharifi E, Boland J. Policy recommendations to increase urban heat stress resilience. Urban Clim. 2018;25:51-63.

Hasan F, Marsia S, Patel K, Agrawal P, Razzak JA. Effective community-based interventions for the prevention and management of heat-related illnesses: a scoping review. Int J Environ Res Public Health. 2021;18(16):8362.

Tomlinson CJ, Chapman L, Thornes JE, Baker CJ. Including the urban heat island in spatial heat health risk assessment strategies: A case study for Birmingham, UK. Int J Health Geogr. 2011;10(1):42. doi:10.1186/1476-072X-10-42.

Brian [cited 2025 Jun 05]. Available from: https://www.proquest.com/docview/229619663?pq-origsite=gscholar&fromopenview=true&sourcetype=Scholarly%20Journals.

**Low and Middle Income Countries (LMIC) excluding India**

McElroy S, Ilango S, Dimitrova A, Gershunov A, Benmarhnia T. Extreme heat, preterm birth, and stillbirth: a global analysis across 14 lower-middle income countries. Environ Int. 2022;158:106902.

Lakhani S, Ambreen S, Padhani ZA, Fahim Y, Qamar S, Meherali S, Lassi ZS. Impact of ambient heat exposure on pregnancy outcomes in low-and middle-income countries: A systematic review. Women's Health. 2024;20:17455057241291271.

Siddik MAZ, Rahman M. Trend analysis of maximum, minimum, and average temperatures in Bangladesh: 1961–2008. Theor Appl Climatol. 2014;116:721-30.

Cheng J, Xu Z, Zhu R, Wang X, Jin L, Song J, Su H. Impact of diurnal temperature range on human health: a systematic review. Int J Biometeorol. 2014;58:2011-24. [Meta Analysis]

McMahon K, Baylis K, Sweeney S, Funk C. Does humidity matter? Prenatal heat and child health in South Asia.

Green H, Bailey J, Schwarz L, Vanos J, Ebi K, Benmarhnia T. Impact of heat on mortality and morbidity in low and middle income countries: a review of the epidemiological evidence and considerations for future research. Environ Res. 2019;171:80-91.

Sapari H, Selamat MI, Isa MR, Ismail R, Mahiyuddin WRW. The impact of heat waves on health care services in low-or middle-income countries: protocol for a systematic review. JMIR Res Protoc. 2023;12(1):e44702.

Haque F, Lampe FC, Hajat S, Stavrianaki K, Hasan ST, Faruque AS, et al. Is heat wave a predictor of diarrhoea in Dhaka, Bangladesh? A time-series analysis in a South Asian tropical monsoon climate. PLoS Glob Public Health. 2024;4(9):e0003629.

Ioannou LG, Foster J, Morris NB, Piil JF, Havenith G, Mekjavic IB, et al. Occupational heat strain in outdoor workers: a comprehensive review and meta-analysis. Temperature. 2022;9(1):67-102. [Meta Analysis]

Habibi P, Razmjouei J, Moradi A, Mahdavi F, Fallah-Aliabadi S, Heydari A. Climate change and heat stress resilient outdoor workers: findings from systematic literature review. BMC Public Health. 2024;24(1):1711.

Samuelson H, Baniassadi A, Lin A, González PI, Brawley T, Narula T. Housing as a critical determinant of heat vulnerability and health. Sci Total Environ. 2020;720:137296. [SIMULATION BASED]

Tham S, Thompson R, Landeg O, Murray KA, Waite T. Indoor temperature and health: a global systematic review. Public Health. 2020;179:9-17.

Teare J, Mathee A, Naicker N, Swanepoel C, Kapwata, Balakrishna Y, et al. Dwelling characteristics influence indoor temperature and may pose health threats in LMICs. Ann Glob Health. 2020;86(1):91.

McMichael AJ, Wilkinson P, Kovats RS, Pattenden S, Hajat S, Armstrong B, et al. International study of temperature, heat and urban mortality: the 'ISOTHURM'project. Int J Epidemiol. 2008;37(5):1121-31.

Basu R. High ambient temperature and mortality: A review of epidemiologic studies from 2001 to 2008. Environ Health. 2009;8(40). doi:10.1186/1476-069X-8-40.

Bell ML, O'neill MS, Ranjit N, Borja-Aburto VH, Cifuentes LA, Gouveia NC. Vulnerability to heat-related mortality in Latin America: a case-crossover study in Sao Paulo, Brazil, Santiago, Chile and Mexico City, Mexico. Int J Epidemiol. 2008;37(4):796-804.

Gouveia N, Hajat S, Armstrong B. Socioeconomic differentials in the temperature–mortality relationship in São Paulo, Brazil. Int J Epidemiol. 2003;32(3):390-7.

Yu W, Mengersen K, Wang X, Ye X, Guo Y, Tong S. Daily average temperature and mortality among the elderly: A meta-analysis and systematic review of epidemiological evidence. Int J Biometeorol. 2012;56(3):569–81. doi:10.1007/s00484-011-0497-3.

Hall JE, Hall ME. Guyton and Hall Textbook of Medical Physiology. 2021. [Physiological Evidence]

Hospers L, Dillon GA, McLachlan AJ, Alexander LM, Kenney WL, Capon A, et al. The effect of prescription and over-the-counter medications on core temperature in adults during heat stress: a systematic review and meta-analysis. EClinicalMedicine. 2024;77.

Campbell S, Remenyi TA, White CJ, Johnston FH. Heatwave and health impact research: A global review. Health Place. 2018;53:210-8.

Green H, Bailey J, Schwarz L, Vanos J, Ebi K, Benmarhnia T. Impact of heat on mortality and morbidity in low and middle income countries: a review of the epidemiological evidence and considerations for future research. Environ Res. 2019;171:80-91.

Razzak JA, Agrawal P, Chand Z, Quraishy S, Ghaffar A, Hyder AA. Impact of community education on heat-related health outcomes and heat literacy among low-income communities in Karachi, Pakistan: a randomised controlled trial. BMJ Glob Health. 2022;7(1):e006845.

Chakraborty T, Hsu A, Manya D, Sheriff G. Disproportionately higher exposure to urban heat in lower-income neighborhoods: a multi-city perspective. Environ Res Lett. 2019;14(10):105003.

Kajjoba D, Kasedde H, Olupot PW, Lwanyaga JD. Evaluation of thermal comfort and air quality of low-income housing in Kampala City, Uganda. Energy Built Environ. 2022;3(4):508-24.

Tawsif S, Alam MS, Al-Maruf A. How households adapt to heat wave for livable habitat? A case of medium-sized city in Bangladesh. Curr Res Environ Sustain. 2022;4:100159.

Hasan F, Marsia S, Patel K, Agrawal P, Razzak JA. Effective community-based interventions for the prevention and management of heat-related illnesses: a scoping review. Int J Environ Res Public Health. 2021;18(16):8362. [Scoping Review]

Cheng Q, Sha S. Resisting the heat wave: Revealing inequalities in matching between heat exposure risk and healthcare services in a megacity. Appl Geogr. 2024;167:103291.

Sapari H, Selamat MI, Isa MR, Ismail R, Mahiyuddin WRW. The impact of heat waves on health care services in low-or middle-income countries: protocol for a systematic review. JMIR Res Protoc. 2023;12(1):e44702. [Systemetic Review]

Pritipadmaja, Garg RD, Sharma AK. Assessing the cooling effect of blue-green spaces: implications for Urban Heat Island mitigation. Water. 2023;15(16):2983.

Kumar D, Shekhar S. Spatial distribution analysis of urban blue-green spaces for mitigating excessive heat with earth observation systems. Environ Challenges. 2021;5:100390.

Hunter RF, Nieuwenhuijsen M, Fabian C, Murphy N, O'Hara K, Rappe E, et al. Advancing urban green and blue space contributions to public health. Lancet Public Health. 2023;8(9):e735-e742.

Hosen MI, Hasan MM, Talha MD, Akter MM, Nasher NR. Exploring the cooling benefits of Urban Lakes: A multi-year analysis of Dhaka, Bangladesh. HydroResearch. 2025.

Bhan SC, Kumar P, Paul S. Effect of Temperature on Gender-Specific All-Cause Mortality: A Study of the City in Northern India. Clim Health J. 2022;2(1).

Lakhoo DP, Brink N, Radebe L, Craig MH, Pham MD, Haghighi MM, et al. A systematic review and meta-analysis of heat exposure impacts on maternal, fetal and neonatal health. Nat Med. 2025;31(2):684-94.

Bell ML, O'neill MS, Ranjit N, Borja-Aburto VH, Cifuentes LA, Gouveia NC. Vulnerability to heat-related mortality in Latin America: a case-crossover study in Sao Paulo, Brazil, Santiago, Chile and Mexico City, Mexico. Int J Epidemiol. 2008;37(4):796-804.

Zhu Y, He C, Bell M, Zhang Y, Fatmi Z, Zhang Y, et al. Association of ambient temperature with the prevalence of intimate partner violence among partnered women in low-and middle-income South Asian countries. JAMA Psychiatry. 2023;80(9):952-61.

Lusambili A, Filippi V, Nakstad B, Natukunda J, Birch CE, Marsham JH, et al. Community perspectives of heat and weather warnings for pregnant and postpartum women in Kilifi, Kenya. PLoS One. 2024;19(11):e0313781.

Bethancourt HJ, Swanson ZS, Nzunza R, Huanca T, Conde E, Kenney WL, et al. Hydration in relation to water insecurity, heat index, and lactation status in two small-scale populations in hot-humid and hot-arid environments. Am J Hum Biol. 2021;33(1):e23447.

Sampson NR, Gronlund CJ, Buxton MA, Catalano L, White-Newsome JL, Conlon KC, et al. Staying cool in a changing climate: Reaching vulnerable populations during heat events. Glob Environ Change. 2013;23(2):475-84.

RIMES. News_HeatwavePortal_Bangladesh. 2024. Available from: https://rimes.int/News_HeatwavePortal_Bangladesh.

Bunker A, Compoaré G, Sewe MO, Laurent JGC, Zabré P, Boudo V, et al. The effects of cool roofs on health, environmental, and economic outcomes in rural Africa: study protocol for a community-based cluster randomized controlled trial. Trials. 2024;25(1):59.

Montaldo P, Pauliah SS, Lally PJ, Olson L, Thayyil S. Cooling in a low-resource environment: lost in translation. Semin Fetal Neonatal Med. 2015;20(2):72-9.

Shahrujjaman SM, Sikder BB, Zahid D, Pal B. Heat Wave Adaptation Strategies among Informal Workers in an Urban Setting: A Study in Dhaka City, Bangladesh. Nat Hazards Res. 2025.

Johar H, Abdulsalam FI, Guo Y, Baernighausen T, Jahan NK, Watterson J, et al. Community-based heat adaptation interventions for improving heat literacy, behaviours, and health outcomes: a systematic review. Lancet Planet Health. 2025. [Systematic Review]

Quinn C, Quintana A, Blaine T, Chandra A, Epanchin P, Pitter S, et al. Linking science and action to improve public health capacity for climate preparedness in lower-and middle-income countries. Clim Policy. 2022;22(9-10):1146-54.

Jahan I. Urban Heat Island (UHI) Development and Mitigation Measures in Three Bangladesh Cities: Dhaka, Chattogram, and Sylhet [Doctoral dissertation]. University of Delaware; 2024.

Hanif A, Nasar-u-Minallah M, Zia S, Ashraf I. Mapping and analyzing the park cooling intensity in mitigation of urban heat island effect in Lahore, Pakistan. Korean J Remote Sens. 2022;38(1):127-37.

**India**

Rathi SK, Sodani PR, Joshi S. Summer temperature and all-cause mortality from 2006 to 2015 for smart city Jaipur, India. J Health Manage. 2021;23(2):294-301.

de Bont J, Nori-Sarma A, Stafoggia M, Banerjee T, Ingole V, Jaganathan S, et al. Impact of heatwaves on all-cause mortality in India: A comprehensive multi-city study. Environ Int. 2024;184:108461.

Burgess R, Deschenes O, Donaldson D, Greenstone M. The unequal effects of weather and climate change: Evidence from mortality in India. Cambridge, United States: Massachusetts Institute of Technology, Department of Economics. Manuscript. 2014.

Azhar GS, Mavalankar D, Nori-Sarma A, Rajiva A, Dutta P, Jaiswal A, et al. Heat-related mortality in India: excess all-cause mortality associated with the 2010 Ahmedabad heat wave. PLoS One. 2014;9(3):e91831.

Jaswal AK, Padmakumari B, Kumar N, Kore PA. Increasing trend in temperature and moisture induced heat index and its effect on human health in climate change scenario over the Indian sub-continent. J Clim Change. 2017;3(1):11-25.

Mall RK, Chaturvedi M, Singh N, Bhatla R, Singh RS, Gupta A, Niyogi D. Evidence of asymmetric change in diurnal temperature range in recent decades over different agro-climatic zones of India. Int J Climatol. 2021;41(4):2597-2610.

Fritz M. Beyond the heat: The mental health toll of temperature and humidity in India. arXiv preprint arXiv:2503.08761. 2025.

Maurya HK, Joshi N, Suryavanshi S. Projected changes in the heatwave's characteristics and associated population exposure over India under 1.5–3° C warming levels. Stoch Environ Res Risk Assess. 2024;38(7):2521-38.

Singh S, Mall RK, Singh N. Changing spatio-temporal trends of heat wave and severe heat wave events over India: An emerging health hazard. Int J Climatol. 2021;41:E1831-E1845.

Saunik S, Shaw R. Impacts of Climate Change and Heat Waves on Public Health and Livelihoods in India: A Review of Recent Literatures. Available at SSRN 4953783.

Thakkar V, Srinivas V, Siddhappanavara PM, Madappa T, Jeganathan A, Murthy IK. Heatwave health risk index for Karnataka, India. J Clim Change Health. 2025;22:100428.

Ioannou LG, Foster J, Morris NB, Piil JF, Havenith G, Mekjavic IB, et al. Occupational heat strain in outdoor workers: a comprehensive review and meta-analysis. Temperature. 2022;9(1):67-102. [Meta Analysis]

Habibi P, Razmjouei J, Moradi A, Mahdavi F, Fallah-Aliabadi S, Heydari A. Climate change and heat stress resilient outdoor workers: findings from systematic literature review. BMC Public Health. 2024;24(1):1711.

Sahu S, Sett M, Kjellstrom T. Heat Exposure, Cardiovascular Stress and Work Productivity in Rice Harvesters in India: Implications for a Climate Change Future. Ind Health. 2013. doi:10.2486/indhealth.2013-0006.

Patel H, Rao NM, Saha A. Heat exposure effects among firefighters. Case Report. Indian J Occup Environ Med. 2006;10(3):121–3. doi:10.4103/0019-5278.29572.

Khetan AK, Yakkali S, Dholakia HH, Hejjaji V. Adaptation to heat stress: a qualitative study from Eastern India. Environ Res Lett. 2024;19(4):044035.

Samuel DL, Dharmasastha K, Nagendra SS, Maiya MP. Thermal comfort in traditional buildings composed of local and modern construction materials. Int J Sustain Built Environ. 2017;6(2):463-75.

Manu S, Brager G, Rawal R, Geronazzo A, Kumar D. Performance evaluation of climate responsive buildings in India-Case studies from cooling dominated climate zones. Build Environ. 2019;148:136-56.

Srikonda R, Dokiparty BR. Integration of natural ventilation by thermal buoyancy in solar thermal modelling for conservation of thermal energy in vernacular building in India. In: 2010 International Conference on Environmental Engineering and Applications; 2010 Sep; pp. 291-4. IEEE.

Mukhopadhyay B, Weitz CA, Das K. Indoor heat conditions measured in urban slum and rural village housing in West Bengal, India. Build Environ. 2021;191:107567.

McMichael AJ, Wilkinson P, Kovats RS, Pattenden S, Hajat S, Armstrong B, et al. International study of temperature, heat and urban mortality: the 'ISOTHURM'project. Int J Epidemiol. 2008;37(5):1121-31.

Samuelson H, Baniassadi A, Lin A, González PI, Brawley T, Narula T. Housing as a critical determinant of heat vulnerability and health. Sci Total Environ. 2020;720:137296. [SIMULATION BASED]

Weitz CA, Mukhopadhyay B, Das K. Individually experienced heat stress among elderly residents of an urban slum and rural village in India. Int J Biometeorol. 2022;66(6):1145-62.

Mukhopadhyay B, Weitz CA. Heat exposure, heat-related symptoms and coping strategies among elderly residents of urban slums and rural vilages in West Bengal, India. Int J Environ Res Public Health. 2022;19(19):12446.

Agarwala S, Ghoshb P, Scarazzatoc F, Waltld SR. The Association between Adverse Temperature Shocks and Schooling Outcomes in India: Impact Quantification and Mitigation Potentials. Education. 2020;1.

Lala B, Hagishima A. Impact of escalating heat waves on students' well-being and overall health: a survey of primary school teachers. Climate. 2023;11(6):126.

Shrikhande S, Pedder H, Roosli M, Dalvie MA, Ravivarman L, Gasparrini A, et al. Temperature and cardiovascular diseases: exploring associations in India and public health insights. Eur J Public Health. 2022;32(Suppl_3):ckac131-157.

Venugopal V, Chinnadurai J, Lucas R, Vishwanathan V, Rajiva A, Kjellstrom T. The social implications of occupational heat stress on migrant workers engaged in public construction: a case study from southern India. Int J Construct Environ. 2016;7(2):25.

Varghese GM, John G, Thomas K, Abraham OC, Mathai D. Predictors of multi-organ dysfunction in heatstroke. Emerg Med J. 2005;22(3):185-7.

Nanda L, Chakraborty S, Mishra SK, Dutta A, Rathi SK. Characteristics of households' vulnerability to extreme heat: an analytical cross-sectional study from India. Int J Environ Res Public Health. 2022;19(22):15334.

Das S. Effects of Climate Change and Heat Waves on Low Income Urban Workers: Evidence from India. In: Delgado-Ramos GC, editor. Inequality and Climate Change: Perspectives from the South. 2015.

Tran KV, Azhar GS, Nair R, Knowlton K, Jaiswal A, Sheffield P, et al. A cross-sectional, randomized cluster sample survey of household vulnerability to extreme heat among slum dwellers in Ahmedabad, India. Int J Environ Res Public Health. 2013;10(6):2515-43.

Somanathan E, Somanathan R, Sudarshan A, Tewari M. The impact of temperature on productivity and labor supply: Evidence from Indian manufacturing. J Polit Econ. 2021;129(6):1797-1827.

Pradyumna A, Bendapudi R, Zade D, D'Souza M, Tasgaonkar P. Managing the increasing heat stress in rural areas. In: Handbook of Climate Change Resilience. 2018;1-22.

Trahan A, Walshe R, Mehta V. Extreme heat, gender, and access to preparedness measures: An analysis of the heatwave early warning system in Ahmedabad, India. Int J Disaster Risk Reduct. 2023;99:104080.

Dasgupta P, Dayal V, Dasgupta R, Ebi KL, Heaviside C, Joe W, et al. Responding to heat-related health risks: the urgency of an equipoise between emergency and equity. Lancet Planet Health. 2024;8(11):e933-e936.

Venugopal V, Shanmugam R, Johnson P, Lucas R, Jakobsson K. Heat stress and inadequate toilet access at work places in India–a potential hazard to working women in a changing climate. Climanosco Res Articles. 2019.

Golechha M, Mavalankar D, Bhan SC. India: heat wave and action plan implementation in Indian cities. In: Urban Climate Science for Planning Healthy Cities. 2021;285-308.

Debnath KB, Wang X, Peters T, Menon S, Awate S, Patwardhan G, et al. Rural cooling needs assessment towards designing community cooling hubs: case studies from Maharashtra, India. Sustainability. 2021;13(10):5595.

Vellingiri S, Dutta P, Singh S, Sathish LM, Pingle S, Brahmbhatt B. Combating climate change-induced heat stress: assessing cool roofs and its impact on the indoor ambient temperature of the households in the urban slums of Ahmedabad. Indian J Occup Environ Med. 2020;24(1):25-9.

Chandrashekhar V. Heat-proofing India. Science. 2023;381(6665):1393-7.

Arumugam R, Garg V, Mathur J, Reddy N, Gandhi J, Fischer ML. Experimental determination of comfort benefits from cool-roof application to an un-conditioned building in India. Adv Build Energy Res. 2014;8(1):14-27.

Das S, Smith SC. Awareness as an adaptation strategy for reducing mortality from heat waves: evidence from a disaster risk management program in India. Clim Change Econ. 2012;3(02):1250010.

Maheshwari V. Analysis of Public Awareness, Health Risks, and Coping Strategies Against Heat Waves in NCT of Delhi, India. In: Challenges of Disasters in Asia: Vulnerability, Adaptation and Resilience. Singapore: Springer Nature Singapore; 2022;299-324.

Das S. Television is more effective in bringing behavioral change: Evidence from heat-wave awareness campaign in India. World Dev. 2016;88:107-21.

Singh C, Vyas D, Patil S, Ranjit N, Poonacha P, Surampally S. How are Indian cities adapting to extreme heat? Insights on heat risk governance and incremental adaptation from ten urban Heat Action Plans. PLoS Clim. 2024;3(11):e0000484.

Directorate of Environment and Climate Change (DoECC), State Action Plan on Climate Change.

Dwivedi A, Mohan BK. Impact of green roof on micro climate to reduce Urban Heat Island. Remote Sens Appl Soc Environ. 2018;10:56-69.

Khare VR, Vajpai A, Gupta D. A big picture of urban heat island mitigation strategies and recommendation for India. Urban Clim. 2021;37:100845.
